# Neutralizing antibody responses to SARS-CoV-2: a population based seroepidemiological analysis in Delhi, India

**DOI:** 10.1101/2021.12.28.21268472

**Authors:** Pragya Sharma, Ekta Gupta, Saurav Basu, Reshu Aggarwal, Suruchi Mishra, Pratibha Kale, Nutan Mundeja, B S Charan, Gautam Kumar Singh, Mongjam Meghachandra Singh

## Abstract

We conducted this study to estimate seroprevalence of neutralizing antibodies in the general population and to further correlate it with the IgG SARS-CoV-2 IgG levels. This present cross-sectional analysis was conducted as a sequel to a state level community-based seroepidemiological study in Delhi, India. A total of 2564 seropositive samples were selected from 25622 seropositive samples through simple random sampling. Neutralizing capacity was estimated by performing a surrogate virus neutralization test with the sVNT (GenScript) assay. Neutralizing antibody against the SARS-CoV-2 virus was operationally considered as detected when the signal inhibition was ≥30%.

A total of 2233 (87.1%, 95% C.I. 85.7, 88.3) of the 2564 SARS-CoV-2 seropositive samples had detectable neutralizing antibodies. On bi-variate analysis but not on adjusted analysis, Covid-19 vaccination showed a statistically significant association with the presence of neutralizing antibodies (p<0.001). The signal/ cut off (S/CO) of SARS-CoV-2 IgG ranged from 1.00 to 22.8 (median 11.40). In samples with S/CO ≥4.00, the neutralizing antibodies ranged from 94.5 to 100%, while in samples with S/CO <4.00, it ranged from 52.0 to 79.2%. The neutralizing antibody seroprevalence strongly correlated with the S/CO range (r=0.62, p=0.002). In conclusion, in populations with high SARS-CoV-2 seroprevalence, neutralizing antibodies are generated in nearly 9 of 10 seropositive individuals.

## INTRODUCTION

The severe acute respiratory syndrome coronavirus 2 (SARS-CoV-2) has caused a global pandemic with enormous morbidity, mortality, and economic losses. Till date, India has recorded 34,633,255 cases and 473,326 deaths [1].

Protection from SARS-CoV-2 infection is accorded through previous infection or vaccination that substantially reduces the risk of symptomatic and severe disease and that of reinfection [2, 3]. Neutralizing SARS-CoV-2 antibodies once produced are likely to provide durable and effective protection at-least several ensuing months [4].

Several population-based SARS-CoV-2 seroepidemiological studies have been conducted in India. However, these studies primarily focussed on screening antibodies in the participants through antibodies recognizing SARS-CoV-2 spike antigens and/or nucleoprotein [5, 6]. Consequently, it is important to identify the extent to which these populations have also developed protective neutralization antibodies.

This study was therefore conducted with the objective of estimating the seroprevalence of neutralizing antibodies in the general population and to further correlate it with the IgG SARS-CoV-2 IgG levels.

## METHODS

This present cross-sectional analysis was conducted as a sequel to a state level community-based seroepidemiological study which estimated the seroprevalence of IgG SARS-CoV-2 in Delhi, India during August-September 2021. A total of 25,622 (89.6%) of the 28,602 samples had detectable IgG antibodies. The VITROS® assay on VITROS® 3600 (Ortho Clinical Diagnostics, Raritan, NJ, USA) based on chemiluminescent technology having 90% sensitivity and 100% specificity was used for this purpose [7]. The detailed methodology and sampling strategy of the study are being reported elsewhere (preprint).

We conducted screening for neutralizing antibodies against the SARS-CoV-2 virus in 10% of the seropositive samples (n=2564) representing the entire range of signal to cut off (S/CO). The samples for screening were selected through computer-based simple random sampling method. This sample size was adequate at 95% confidence levels with 1.2% absolute precision levels.

Neutralizing capacity was estimated by performing a surrogate virus neutralization test (sVNT) (GenScript, Piscataway, NJ, USA) as per the manufacturers’ instructions. The sVNT kit detects circulating neutralizing anti SARS-CoV-2 antibodies through immune system response either after COVID-19 infection or vaccination. These antibodies prevent the interaction between the ACE2 human cell surface receptor and the receptor binding domain (RBD) of the SARS-CoV-2 spike glycoprotein [8]. Neutralizing antibody to SARS-CoV-2 was operationally considered as detected when the signal inhibition was ≥30%.

Data were analysed with IBM SPSS Statistics for Windows, Version 25.0. Armonk, NY: IBM Corp. A p-value < 0.05 was considered statistically significant The study was approved by the Institutional Ethics Committee, Maulana Azad Medical College & Associated Hospitals, New Delhi vide F.1/IEC/MAMC/85/03/2021/No428 dated 21.08.2021. Electronic and informed consent was obtained from all the study participants.

## RESULTS AND DISCUSSION

A total of 2233 (87.1%, 95% C.I. 85.7, 88.3) of the 2564 SARS-CoV-2 seropositive samples were observed to be having detectable neutralizing antibodies. On bi-variate analysis but not on adjusted analysis, Covid-19 vaccination showed a statistically significant association with the presence of neutralizing antibodies (p<0.001) (Table 1).

**Table 1.** Factors associated with positive neutralization antibodies (N=2564)

| Variable | Total* | Neutralizing Ab present | Adjusted odds | p-value |
| --- | --- | --- | --- | --- |
| Age |  |  |  |  |
| <18 | 487 | 403 (82.8) | 1.1 (0.77, 1.57) | 0.59 |
| ≥18 | 2077 | 1830 (81.1) | 1 |  |
| Sex |  |  |  |  |
| Male | 1093 | 944 (86.4) | 1 | 0.84 |
| Female | 1467 | 1286 (87.7) | 1 (0.78, 1.3) |  |
| History Covid-19 |  |  |  |  |
| Present | 559 | 495 (88.6) | 1.1 (0.83, 1.5) | 0.40 |
| Absent | 1300 | 1117 (85.9) | 1 |  |
| HTN |  |  |  |  |
| Yes | 173 | 148 (85.5) | 1 | 0.35 |
| No | 1686 | 1464 (86.8) | 1.2 (0.77, 2.1) |  |
| DM |  |  |  |  |
| Yes | 119 | 101 (84.9) | 1 | 0.58 |
| No | 1740 | 1511 (86.8) | 1.2 (0.66, 2.1) |  |
| Vaccine doses |  |  |  |  |
| 0 dose | 929 | 769 (82.8) | 1 | 0.09 |
| 1 dose | 430 | 400 (93.0) | 2.1 (0.7, 6.0) |  |
| 2 doses | 500 | 443 (88.6) | 1.7 (0.5, 3.2) |  |
| Vaccine type |  |  |  |  |
| BBV152 | 185 | 164 (88.6) | 1 | 0.49 |
| CHADOX1 NCOV-19 | 742 | 676 (91.1) | 1.2 (0.7, 2.0) |  |

\*Not equal to 2564 due to missing questionnaire data

The signal/ cut off (S/CO) of SARS-CoV-2 IgG ranged from 1.00 to 22.8 (median 11.40). Among the samples with S/CO ≥4.00, the prevalence of neutralizing antibodies ranged from 94.5 to 100%, while in samples with S/CO <4.00, it ranged from 52.0 to 79.2% (Table 2). The neutralizing antibody seroprevalence strongly correlated with the S/CO range (r=0.62, p=0.002)

**Table 2.** S/CO Range and neutralization antibody positivity

| S/CO range | Total samples tested | Total samples positive for neutralization antibody | Percentage of samples positive for neutralization antibody (%) |
| --- | --- | --- | --- |
| 1.00 to 1.99 | 446 | 232 | 52 |
| 2.00 to 2.99 | 309 | 237 | 76.7 |
| 3.00 to 3.99 | 111 | 88 | 79.2 |
| 4.00 to 4.99 | 109 | 103 | 94.5 |
| 5.00 to 5.99 | 109 | 105 | 96.3 |
| 6.00 to 6.99 | 110 | 105 | 95.5 |
| 7.00 to 7.99 | 110 | 109 | 99 |
| 8.00 to 8.99 | 110 | 109 | 99 |
| 9.00 to 9.99 | 110 | 110 | 100 |
| 10.00 to 10.99 | 111 | 111 | 100 |
| 11.00 to 11.99 | 110 | 109 | 99 |
| 12.00 to 12.99 | 111 | 111 | 100 |
| 13.00 to 13.99 | 110 | 109 | 99 |
| 14.00 to 14.99 | 111 | 111 | 100 |
| 15.00 to 15.99 | 110 | 109 | 99 |
| 16.00 to 16.99 | 110 | 110 | 100 |
| 17.00 to 17.99 | 111 | 111 | 100 |
| 18.00 to 18.99 | 59 | 59 | 100 |
| 19.00 to 19.99 | 49 | 47 | 96 |
| 20.00 to 20.99 | 20 | 20 | 100 |
| 21.00 to 21.99 | 19 | 19 | 100 |
| 22.00 to 22.8 | 09 | 09 | 100 |
| Total | 2564 | 2233 | 87.1 |

Previous studies have suggested that neutralizing antibodies to SARS-CoV-2 are able to block the viral infection and provide durable immune response despite the likelihood of waning of overall antibody levels. A large-scale study at Mount Sinai hospital, USA among health care workers also found strong correlation of immunoglobulin G antibody responses against the viral spike protein generated in those with natural SARS-CoV-2 infection [9]. Our study results further indicate that in the populations with high SARS-CoV-2 seroprevalence, neutralizing antibodies are generated in nearly 9 of 10 seropositive individuals, with no statistically significant variation in the observed risk based on their age, sex, and comorbidity status.

This study also suggests that a higher signal to cut-off ratio may be considered as an indirect predictor for the presence of neutralizing antibody response. This finding may have a clinical implication towards recommending booster doses in vulnerable populations such as the immunocompromised and healthcare workers.

Certain variants of concern especially the Delta have demonstrated the ability to bypass existing immune response and elicit symptomatic disease even in vaccinated and more rarely in recovered individuals [10]. However, in this study, vaccination with at-least one dose of vaccine, either CHADOX1 NCOV-19 (Covishield) or BBV152 (Covaxin) was likely to induce robust neutralizing antibody response.

## Data Availability

All data produced in the present study are available upon reasonable request to the authors

## Acknowledgments

We thank all the district Nodal officers of Delhi for facilitating the data and sample collection. We express thanks to the ATE Chandra Foundation and ACT grants, and the IDFC Foundation for technical support. We also thank Ms. Arti Kakkar for her assistance with data management for this investigation.

